# Clinical, psychosocial and demographic factors affect decisions in SLE people

**DOI:** 10.1101/2024.03.25.24304643

**Authors:** Pedraza-Meza Luis Miguel, Hernández-Ledesma Ana Laura, Alejandra E Ruiz-Contreras, Medina-Rivera Alejandra, Martínez Domingo

## Abstract

Neurological and psychiatric manifestations affect most lupus individuals and include depression, anxiety, mood disorders, and cognitive dysfunction. Although there is evidence supporting suboptimal decision-making in lupus and its association with glucocorticoids consumption, it is not clear what variables impact such decisions. The aim of this study is to explore how social, clinical, psychological, and demographic factors impact social and temporal decision-making in people with lupus. Through a within-subjects experimental-design, our participants responded to social, clinical, psychological, and demographic electronic questionnaires. Then, they participated in two behavioral economics experiments: the third-party dictator game, and the delay discounting task. Our results show that hostility, and age are essential predictors of social decisions, whereas obsessive-compulsiveness and anxiety better predict temporal decisions. These variables behave as expected, but anxiety shows unexpected results: most anxious people act patiently and prefer delayed but bigger rewards. Finally, clinical factors are critical decision predictors for social and temporal decisions. When people are in remission, they tend to impose higher punishment on those who violate the social norm, and they also tend to prefer immediate rewards. When taking glucocorticoids, they also prefer immediate rewards, and as the dosage of glucocorticoids intake increases, they tend to impose higher punishment on norm violators. Clinicians, researchers, and practitioners must consider the side effects of glucocorticoids on decision-making.

## Introduction

Systemic Lupus Erythematosus (SLE) is an autoimmune disease characterized by a chronic activation of the immune system towards the body’s own cells, causing systemic damage. Neuropsychiatric Systemic Lupus Erythematosus (NPSLE) encompasses both neurological and psychiatric manifestations, and it has been estimated that it affects up to 95% of people with SLE (Aguilera-Pickens & Abud-Mendoza, 2013; Liu et al., 2022; Sarwar et al., 2021).

Pain, fatigue, cognitive dysfunction, major depression, and anxiety prevalence are estimated to be higher among people with SLE than in the general population. Depression and anxiety are considered among the most frequent and impactful neuropsychiatric manifestations of SLE; depression prevalence ranges between 2% and 91.7%, whereas anxiety is estimated to affect between 6.4% and 40% of the people with SLE (Kósa et al., 2022).

The presence of depression and anxiety in people with SLE can be triggered by the integration of social, economic, biological, and psychological factors and has been associated with other manifestations such as fatigue, sleep problems, and cognitive dysfunction(Narupan et al., 2022; Schwartz et al., 2019).

The cognitive dysfunction (CD) prevalence in SLE is estimated to be twice that of the general population, and ranges from 3% to 80%. Besides the presence of depression and anxiety, other factors such as sleep deprivation and fatigue have been proposed to contribute to CD. Its manifestations are highly heterogeneous and can include mood disorders, impairments in attention, working memory, executive functioning, and visuospatial processing (Ho et al., 2018; Seet et al., 2021).

Several mechanisms have been proposed as contributing factors to the development of NPSLE symptoms, including the presence of inflammatory cytokines and autoantibodies, blood-brain barrier disruption, and cerebrovascular alterations; it has also been suggested that NPSLE could be attributed to the damage generated by the disease and the application of different treatments (Kósa et al., 2022; Liu et al., 2022; Sarwar et al., 2021).

In this line, the “standard of care” for SLE management aims to reach remission and involves, mainly, the use of glucocorticoids (GCs) and hydroxychloroquine. Although effective, the use of GCs has also been associated with many adverse effects, such as organ damage and infections (Porta et al., 2020). In SLE, side effects of GCs have been associated with both the use of high doses in a short time and to their chronic consumption; importantly, the risk of organ damage is estimated to increase up to 2.8% per mg of prednisone consumed per day (Mejía-Vilet & Ayoub, 2021).

GCs can potentially exert effects on neurocognitive function, through interactions with neuronal receptors on the prefrontal cortex, the hippocampus and the basolateral amygdala. For instance, previous studies have reported that the use of high doses during a short period of time is associated with deficits in declarative memory; and that a decrease in working memory is observed when high doses of GCs are consumed on a daily basis. Other studies have observed that the use of moderate doses of GCs on a long-term basis is associated with a decline in cognitive flexibility and lower decision-making capability (Montero-López et al., 2016; Seet et al., 2021).

In summary, NPSLE is estimated to affect up to 95% of people with SLE; with depression, anxiety, mood disorders, and cognitive dysfunction among the most frequent manifestations. Although there is evidence supporting the loss of decision-making capacity in lupus and its association with GCs consumption, there is a research gap concerning decision-making in people with SLE. Behavioral economics research has brought insights into social and temporal decision-making. Social decisions have been studied through social punishment tasks. Whereas, temporal decisions have been studied by delay-discounting tasks.

Social punishment is the behavior of incurring personal costs to punish norm violators (Fehr & Gächter, 2002; Rodrigues et al., 2020). When an individual perceives unfair situations, adverse emotional responses emerge (Rodrigues et al., 2018; Seip et al., 2009) leading to punishing norm violations. This behavior has been associated with anger (Rodrigues et al., 2018), impulsiveness (Crockett et al., 2010), and sex (Singer et al., 2006). And it is reported that punishment behavior increases when the impact on norm-violators is high and the cost of punishment is low (Ostrom et al., 1992).

Delay discounting is the process that permits an individual to make value comparisons between immediate versus delayed rewards (Loewenstein, 1988). Numerous studies indicate that delay discounting anomalies are closely linked to poor psychological health, including depression, anxiety, and perceived stress (Campbell & Egede, 2022; Macedo et al., 2022). It is also associated with lower levels of social support, alcohol consumption, and severe depressive disorders (Felton et al., 2020).

The advance in behavioral economics around social and temporal decision-making suggests that many symptoms could affect social and temporal decision-making in people with SLE; however, experimental studies on people with SLE concerning decision-making are scarce. The aim of this study is to explore how social, clinical, psychological, and demographic factors impact social and temporal decision-making in people with SLE.

## Methods

A total of 51 people with SLE were recruited via the Mexican Lupus Registry database (Reyes-Perez et al., 2023), between November 2022 and May 2023. All of them provided informed consent through the REDCap (Research Electronic Data Capture) platform. They were also asked to respond to electronic questionnaires with social, clinical, psychological, and demographic variables. 22 people didn’t complete their responses, so they were excluded from the analysis. We previously piloted for social decisions, revealing a mean true effect size of d = 0.72; thus, our expected statistical power is 76.87%. For temporal decisions, based on literature (Weinsztok et al., 2021), we assumed a mean true effect size of d = 0.82, then expecting a power of 86.62%. This research protocol was approved by the Ethics Research Committee of the Neurobiology Institute at the National Autonomous University of Mexico (protocol-093).

Social variables included the socioeconomic level index and quality of life. The socioeconomic level was calculated following the rule of the Mexican Association of Market and Opinion Intelligence Agencies (Comité de Nivel Socioeconómico AMAI, 2021), whereas the quality of life was computed based on the Spanish version of the World Health Organization Quality of Life Questionnaire (World Health Organization, 2004).

Clinical variables included glucocorticoids consumption and the illness’s remission state. For glucocorticoids consumption, participants were asked to provide a detailed list of medication intake; to confirm the information, all participants were asked to share their medical prescription. Concerning the illness’s remission state, participants were asked if their medical provider informed them that they were in a remission state (reduction or complete disappearance of the symptoms of lupus) or not. In case the participant didn’t know it, the response was coded as unknown.

Psychological variables came from three questionnaires: the Symptom Checklist-90 (SCL-90), the Spielberg’s State and Trait Anxiety Inventory (STAI-S & STAI-T), and the Spielberg’s State and Trait Depression Questionnaire (ST-Dep). SCL-90 included the following variables: anxiety, hostility, paranoid ideation, obsessive-compulsive, depression, phobic anxiety, somatization, and interpersonal sensitivity. STAI-S reported state anxiety, whereas STAI-T reported trait anxiety. Finally, ST-Dep reported the variables of state and trait depression.

Data on demographic variables included the sex, age, and age group of the participants were also registered

Our approach was a within-subjects experimental design. So, once the individuals completed all the questionnaires, they participated in two standard behavioral-economic tasks. The first task was a third-party dictator game, focused on evaluating social punishment behavior in people with SLE. The second task was the temporal discounting reward choice, focused on evaluating delay discounting preferences.

In the third-party dictator game, participants saw a video of two people with SLE playing the dictator game, but instead of tokens, they played with single pieces of chocolates. At the beginning, each player won 7 chocolates by solving math problems. At the end, the dictator decides how to divide the 14 chocolates between herself and the other player (i.e., the recipient); indeed, the dictator played selfishly and decided to keep all the 14 chocolates for herself.

After this point, our participant (the third person) was endowed with 7 chocolates, and we offered her the possibility of punishing the unfair player (i.e., the dictator) in exchange for some of her chocolates. Specifically, we asked our participants: “Would you pay 3 chocolates to punish the unfair dictator by 70% off? 70% off means, take off the 70% of the dictator’s 14 chocolates”. We repeated this essay many times, varying randomly the number of chocolates (i.e., the cost of punishment) between 0 and 7, and also varying the size of the punishment between 0% and 100%, increasing each time by 20%. The responses were recorded as “yes” when participants decided to punish, or “not” when participants decided not to punish. Furthermore, the reaction time, size of punishment, and cost of punishment were recorded.

In the temporal discounting reward choice, our participants were endowed with 7 chocolates. They were advised that this second task was independent of the first one. Once each participant received 7 chocolates, we explained to them that they could keep such an amount or opt for another number of chocolates, if they were able to wait for it. Specifically, we asked them: Do you prefer 7 chocolates today, or 13 chocolates waiting 3 days? We repeated this assay many times, varying each time the waiting time between 1 and 5 days, and varying how many chocolates she would receive after waiting. The received chocolates after waiting varied between 8 and 32, with a gaining rate between 1 and 5. A gaining rate of 1 means that, one chocolate will be added every day of waiting; a gaining rate of 2 means that, two chocolates will be added every day of waiting; and so on. Importantly, the comparison base “7 chocolates today” always remains the same for all the assays.

In this task, the main response was coded either as “immediate reward”, when the participant preferred 7 chocolates today, or as “delayed reward”, when the participant preferred waiting to obtain a bigger reward. The reaction time, the size of the future reward, the waiting time, the comparison base (7 chocolates), and the gaining rate, were recorded.

Once we collected all the data from each participant, our analysis pipeline was divided in three steps. As the first step, we implemented stepwise logistic regression models (backward induction, see below) to identify which dependent variables significantly explain social or temporal decisions. In the second step, we applied a 10-fold cross-validation partition to better estimate the model’s accuracy and identify the most important variables to predict both, the temporal and social decisions. Variables’ importance were assessed by measuring the absolute value of the t-statistics for each validated model parameter, then, normalized from 100, for the most influential, to 0, for the least influential. The accuracy of the model was evaluated using permutation-based scores.

Finally, as a third step, the validated estimated parameters were ranked by their relevance (calculated in the previous step), to identify the most influential variables, which increases the chance that our results can be generalized.

For social decisions, the model included the punishing or not-punishing response as the target variable (i.e., dependent variable), whereas, for predicting variables (i.e., independent variables), the model included: socioeconomic level, quality of life, glucocorticoids consumption (coded as the dose taken or binary consumption), remission, anxiety, hostility, paranoid ideation, obsessive-compulsive, depression, phobic anxiety, somatization, interpersonal sensitivity, state anxiety, trait anxiety, trait depression, sex, age, age group, the reaction time, size of punishment, and cost of punishment.

For temporal decisions, the model included the immediate or waiting reward response as the target variable (i.e., dependent variable), whereas, for predicting variables (i.e., independent variables), the model included: socioeconomic level, quality of life, glucocorticoids consumption (coded as the dose taken or binary consumption), remission, anxiety, hostility, paranoid ideation, obsessive-compulsive, depression, phobic anxiety, somatization, interpersonal sensitivity, state anxiety, trait anxiety, trait depression, sex, age, age group, the reaction time, size of future reward, waiting time, and the gaining rate.

All statistical analysis was implemented in R 4.3.3 (R Core Team, 2024). The scripts and datasets are available on https://github.com/NeuroGenomicsMX/Factors_affecting_decisions_in_SLE, and https://zenodo.org/records/10806272, respectively.

## Results

We evaluated 29 Mexican people with SLE, with a mean age of 34.3 (±7.5) years, ranging from 18 to 48 years. Twenty-seven (93.1%) of the participants were women, with a mean age of 33.8 (±7.6) years, male participants (6.9%) had a mean age of 40 (±1.4) years. Fifteen (51.7%) participants reported consuming glucocorticoids daily as part of their treatment, with the doses ranging from 2.5 to 50 mg per day.

For the third-party dictator game, we recorded 1160 binary responses, 40 from each participant. For the temporal discounting task, we recorded 725 binary responses, 25 from each participant.

The aim of this study was to explore how social, clinical, psychological, and demographic factors impact social and temporal decision-making in Our approach was a within-subjects experimental design. So, once the individuals completed all the questionnaires, they participated in two standard behavioral-economic tasks. The first task was a third-party dictator game, focused on evaluating social punishment behavior in people with SLE. The statistical analysis was divided in three steps: i) logistic regression models to identify variables explaining social or temporal decisions; ii) cross validation assessment of the model accuracy, and variables relevance to predict social and temporal decisions; and iii) identification of the most influential variables in our model.

We built a stepwise logistic regression model (backward induction) to explain social decision-making (p-value < 0.0001), and identified explanatory variables (Table 1). Regarding social factors, only quality of life showed a significant effect on Among the clinical factors, SLE remission and glucocorticoids dose had a significant effect on social decision-making. Within psychological factors, anxiety, depression, hostility, paranoid ideation, somatization, obsessive-compulsive, interpersonal sensitivity, state depression, trait depression, and state-anxiety showed significance on social decision-making; only phobic-anxiety and trait anxiety didn’t show significant effects. For the demographic factors, only age had a significant effect on social decision-making. Finally, from task parameters, reaction time, personal cost, and punishment size exhibited a significant effect on social decision-making.

**Table 1:**
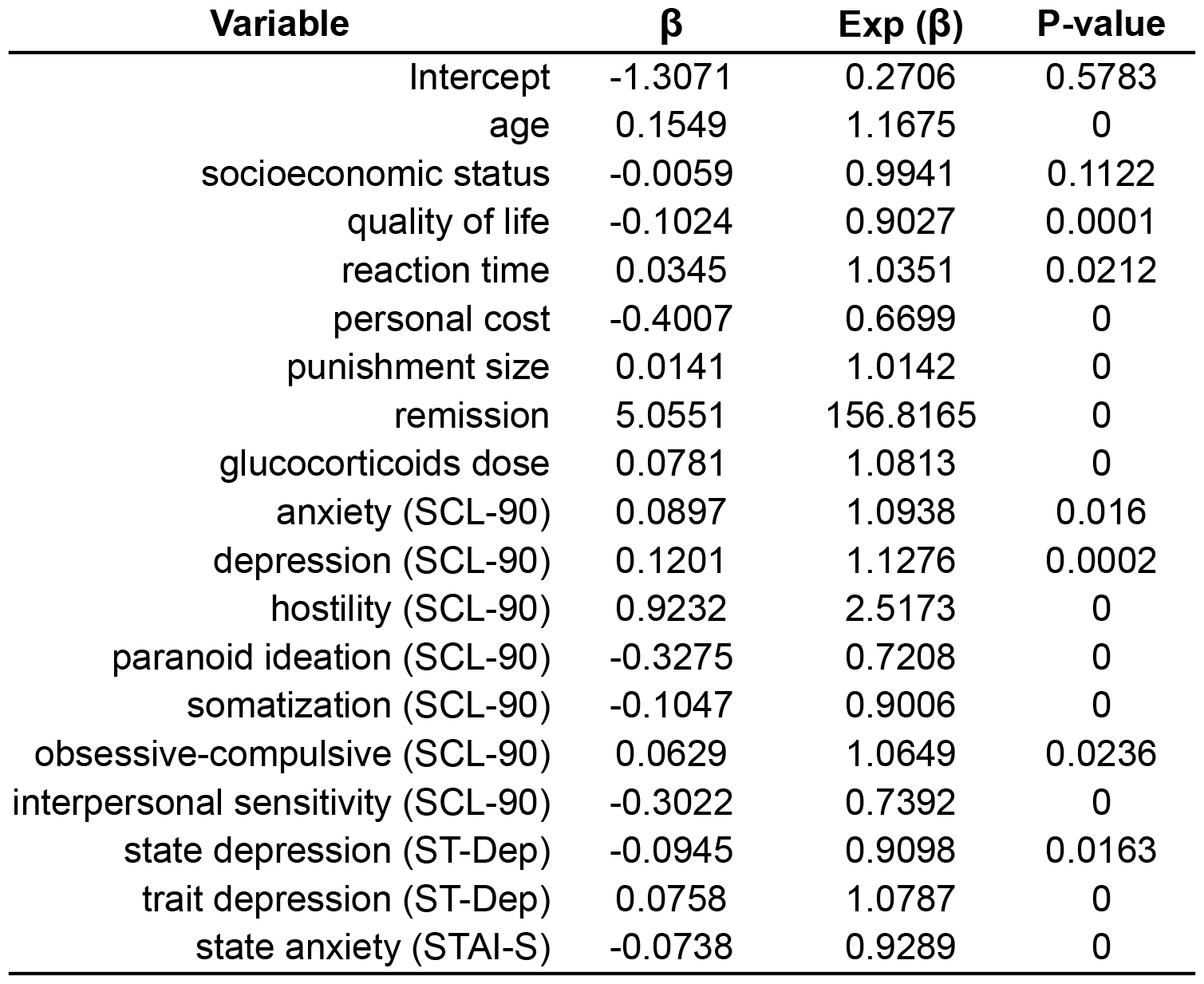
Stepwise logistic regression results for social decisions.

Using the same strategy, we built a model (p-value < 0.0001) to explain temporal decision-making (Table 2). All social factors (i.e., socioeconomic status and quality of life) showed a significant effect on temporal decision-making. In terms of clinical factors, SLE remission and glucocorticoids consumption showed a significant effect. Within psychological factors, anxiety, depression, hostility, paranoid ideation, obsessive-compulsive, interpersonal sensitivity, state depression, trait depression, and state anxiety showed significance on social decision-making; only somatization, phobic-anxiety and trait anxiety didn’t show significant effects. Concerning demographic factors, sex, and age group had a significant effect. Finally, from task parameters, size of the future reward, and the gaining rate, showed significance.

**Table 2:** Stepwise logistic regression results for temporal decisions.

| Variable | $\beta$ | Exp ($\beta$) | P-value |
| --- | --- | --- | --- |
| (Intercept) | 25.1054 | 80008023827.6817 | 0 |
| sex | -2.1887 | 0.1121 | 0 |
| age group | 0.7795 | 2.1804 | 0.0003 |
| socioeconomic status | -0.0486 | 0.9525 | 0 |
| quality of life | -0.3328 | 0.7169 | 0 |
| future reward | 0.039 | 1.0398 | 0.0934 |
| remission | 11.401 | 89409.5809 | 0 |
| gaining rate | -0.6791 | 0.5071 | 0 |
| glucocorticoids consumption | 3.7262 | 41.5192 | 0 |
| anxiety (SCL-90) | -0.9633 | 0.3816 | 0 |
| depression (SCL-90) | 0.4556 | 1.5772 | 0 |
| hostility (SCL-90) | 0.6759 | 1.9658 | 0 |
| paranoid ideation (SCL-90) | -1.1529 | 0.3157 | 0 |
| somatization (SCL-90) | -0.0417 | 0.9591 | 0.0537 |
| obsessive-compulsive (SCL-90) | 0.7108 | 2.0356 | 0 |
| interpersonal sensitivity (SCL-90) | 0.2174 | 1.2429 | 0.0031 |
| state depression (ST-Dep) | -0.3451 | 0.7082 | 0 |
| trait depression (ST-Dep) | -0.1104 | 0.8955 | 0.0001 |
| state anxiety (STAI-S) | 0.1002 | 1.1054 | 0.0001 |

We applied a 10-fold cross-validation partition to identify the most relevant variables for the model explaining social decisions (Figure 1). We observed that for social factors, age was the most influential variable. All clinical factors were influential: glucocorticoids dose, and SLE remission state. Concerning psychological factors, hostility and somatization showed the greatest influence. Regarding demographic factors, only age was significant. Finally, for task parameters, the personal cost (i.e., how many chocolates the participant was willing to pay to punish the unfair dictator) was the most significant variable.

**Figure 1:**
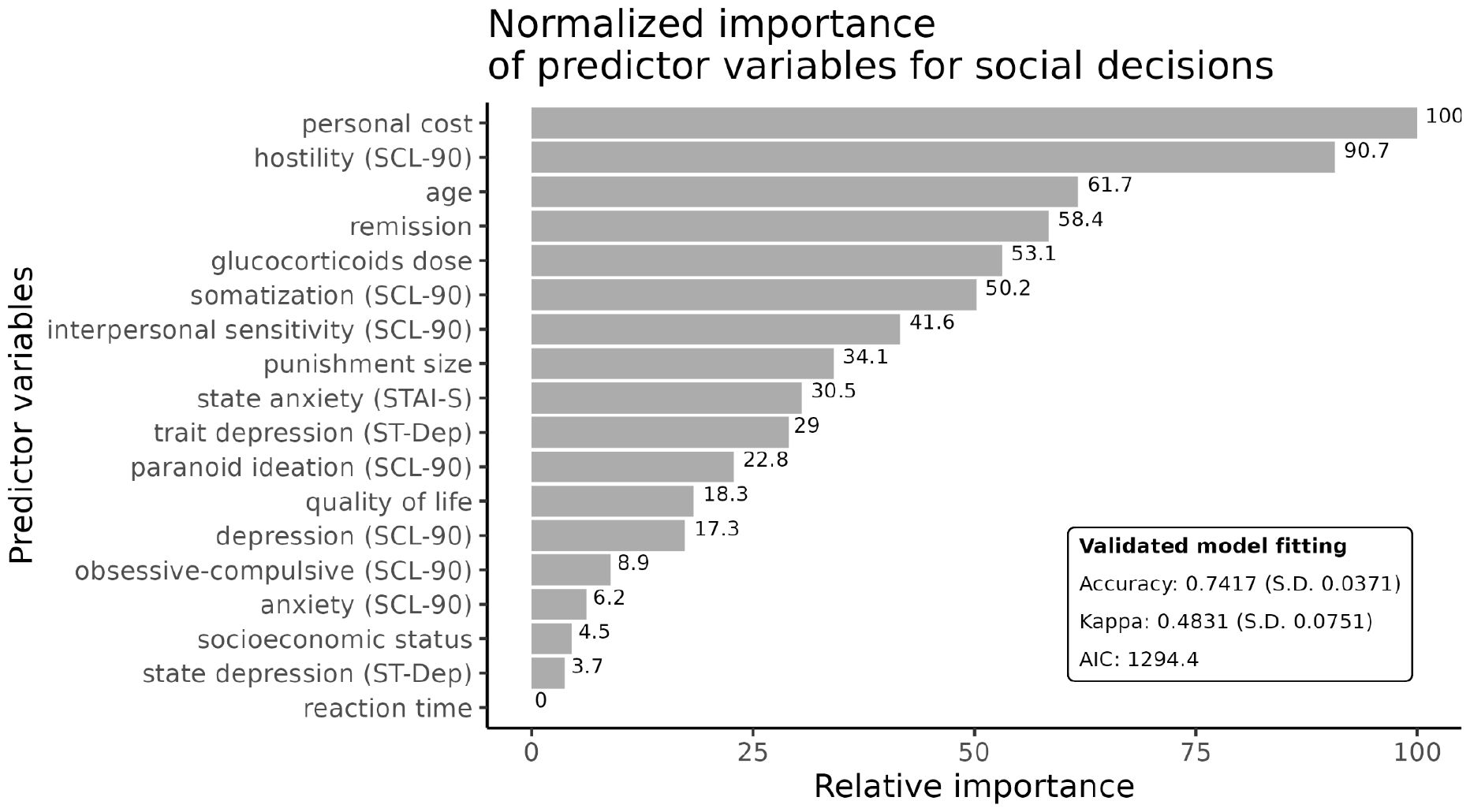
Importance variables for social decisions. X axis shows the relative importance based on a 10 fold cross validation of variables used to build our model to explain social decisions. Y axis shows the predictor variables sorted by relative importance.

We used the same cross validation strategy for the variables used to build the model to explain temporal decisions (Figure 2). Among social factors, quality of life, and socioeconomic status were important predicting variables. Concerning clinical factors, SLE remission state was very influential. For psychological factors, anxiety, obsessive-compulsive, depression, and paranoid ideation showed higher importance. None of the demographic factors were among the most influential variables. Finally, about task parameters, the gaining rate, i.e., the number of chocolates gained by each day of delay, was a critical predictor for temporal decision.

**Figure 2:**
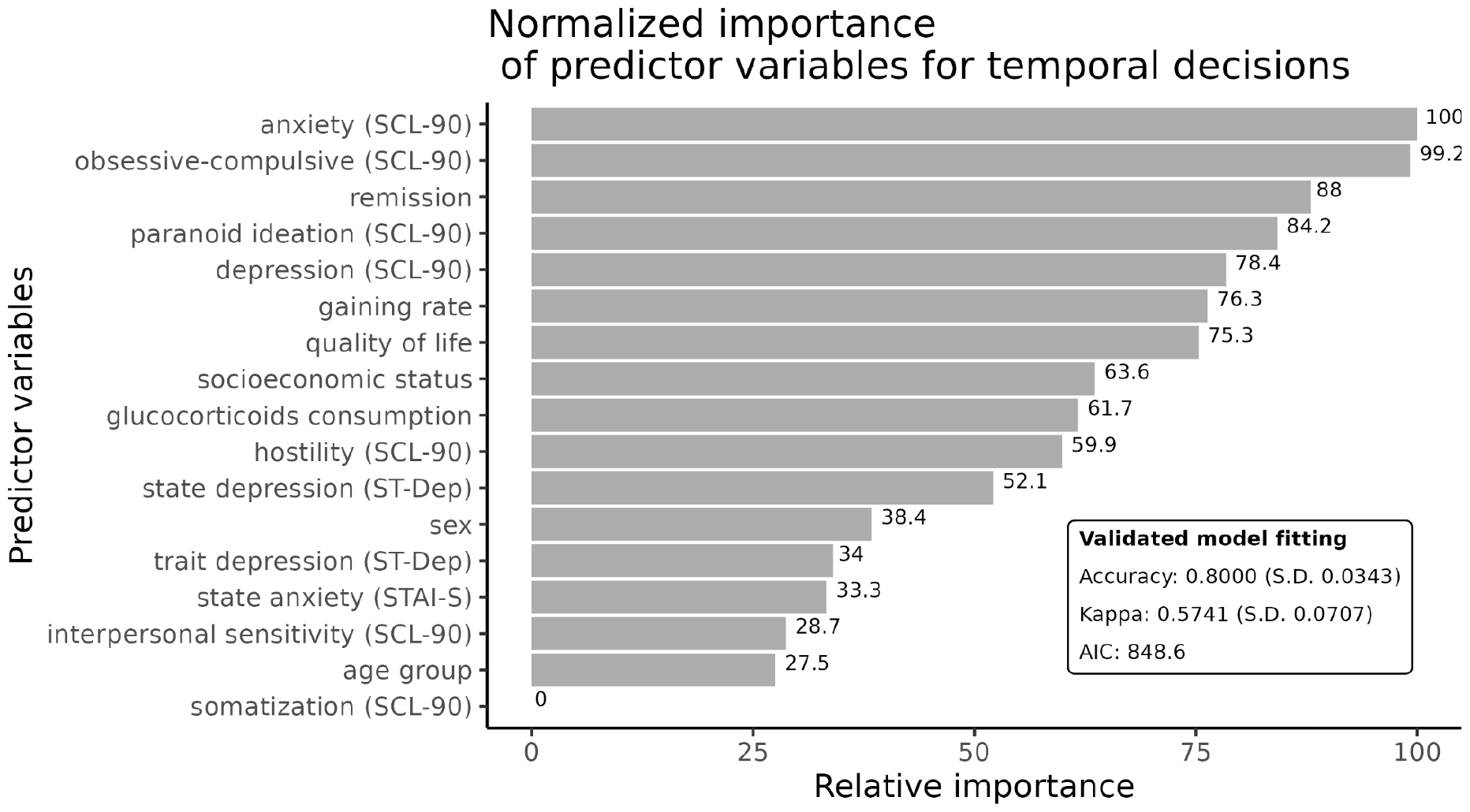
Importance of variables for temporal decision. X axis shows the relative importance based on a 10 fold cross validation of variables used to build our model to explain social decisions. Y axis shows the predictor variables sorted by relative importance.

Once variables were ranked, we estimated the models’ validated parameters that were computed for social (Table 3) and temporal (Table 4) decisions, including percentage odds variation.

**Table 3:** Cross-validated logistic regression results for social decisions, arranged by variables’ importance.

| Variable | $\beta$ | Exp ($\beta$) | Odds % variation | P-value | Importance | Category |
| --- | --- | --- | --- | --- | --- | --- |
| personal cost | -0.4255 | 0.6535 | -34.65 | < 0.0001 | 100 | task related variables |
| hostility (SCL-90) | 0.8991 | 2.4575 | 145.75 | < 0.0001 | 90.7 | psychological variables |
| age | 0.1556 | 1.1684 | 16.84 | < 0.0001 | 61.7 | demographic variables |
| remission | 4.8282 | 124.9806 | 12398.06 | < 0.0001 | 58.4 | clinical variables |
| glucocorticoids dose | 0.0746 | 1.0774 | 7.74 | < 0.0001 | 53.1 | clinical variables |
| somatization (SCL-90) | -0.1049 | 0.9004 | -9.96 | < 0.0001 | 50.2 | psychological variables |
| interpersonal sensitivity (SCL-90) | -0.3072 | 0.7355 | -26.45 | < 0.0001 | 41.6 | psychological variables |
| punishment size | 0.0139 | 1.014 | 1.4 | < 0.0001 | 34.1 | task related variables |
| state anxiety (STAI-S) | -0.0693 | 0.933 | -6.7 | < 0.0001 | 30.5 | psychological variables |
| trait depression (ST-Dep) | 0.0811 | 1.0845 | 8.45 | < 0.0001 | 29 | psychological variables |
| paranoid ideation (SCL-90) | -0.3237 | 0.7235 | -27.65 | < 0.0001 | 22.8 | psychological variables |
| quality of life | -0.0913 | 0.9128 | -8.72 | 0.0001 | 18.3 | social variables |
| depression (SCL-90) | 0.1097 | 1.1159 | 11.59 | 0.0002 | 17.3 | psychological variables |
| obsessive-compulsive (SCL-90) | 0.0751 | 1.078 | 7.8 | 0.0046 | 8.9 | psychological variables |
| anxiety (SCL-90) | 0.0907 | 1.0949 | 9.49 | 0.011 | 6.2 | psychological variables |
| socioeconomic status | -0.0084 | 0.9916 | -0.84 | 0.0179 | 4.5 | social variables |
| state depression (ST-Dep) | -0.0858 | 0.9178 | -8.22 | 0.0224 | 3.7 | psychological variables |
| reaction time | 0.0276 | 1.0279 | 2.79 | 0.059 | 0 | task related variables |

**Table 4:** Cross-validated logistic regression results for temporal decisions, sorted by variables’ importance.

| Variable | $\beta$ | Exp ($\beta$) | Odds % variation | P-value | Importance | Category |
| --- | --- | --- | --- | --- | --- | --- |
| anxiety (SCL-90) | -0.9203 | 0.3984 | -60.16 | < 0.0001 | 100 | psychological variables |
| obsessive-compulsive (SCL-90) | 0.6784 | 1.9708 | 97.08 | < 0.0001 | 99.2 | psychological variables |
| remission | 10.8491 | 51485.2603 | 5148426.03 | < 0.0001 | 88 | clinical variables |
| paranoid ideation (SCL-90) | -1.1148 | 0.328 | -67.2 | < 0.0001 | 84.2 | psychological variables |
| depression (SCL-90) | 0.4108 | 1.508 | 50.8 | < 0.0001 | 78.4 | psychological variables |
| gaining rate | -0.6221 | 0.5368 | -46.32 | < 0.0001 | 76.3 | task related variables |
| quality of life | -0.3178 | 0.7277 | -27.23 | < 0.0001 | 75.3 | social variables |
| socioeconomic status | -0.0452 | 0.9558 | -4.42 | < 0.0001 | 63.6 | social variables |
| glucocorticoids consumption | 3.4771 | 32.3655 | 3136.55 | < 0.0001 | 61.7 | clinical variables |
| hostility (SCL-90) | 0.6406 | 1.8976 | 89.76 | < 0.0001 | 59.9 | psychological variables |
| state depression (ST-Dep) | -0.3339 | 0.7161 | -28.39 | < 0.0001 | 52.1 | psychological variables |
| sex | -2.0208 | 0.1325 | -86.75 | < 0.0001 | 38.4 | demographic variables |
| trait depression (ST-Dep) | -0.1153 | 0.8911 | -10.89 | < 0.0001 | 34 | psychological variables |
| state anxiety (STAI-S) | 0.0946 | 1.0992 | 9.92 | < 0.0001 | 33.3 | psychological variables |
| interpersonal sensitivity (SCL-90) | 0.2464 | 1.2795 | 27.95 | < 0.0001 | 28.7 | psychological variables |
| age group | 0.6807 | 1.9753 | 97.53 | 0.0001 | 27.5 | demographic variables |
| somatization (SCL-90) | -0.0178 | 0.9824 | -1.76 | 0.3065 | 0 | psychological variables |

In the top five of the most influential predicting variables for social decision (Table 3) we see: task parameters, psychological, demographic, and clinical variables. All of them with a significance of <0.0001. The most influential variable is personal cost, which belongs to the parameters of the decision-making task. Specifically, personal cost describes how many chocolates the participants will be willing to pay to punish a norm violator (i.e., the unfair dictator). The results indicate that, the odds of social punishment decreases by 34.65% as the cost increases by one unit (i.e., one chocolate). Hostility is the most influential variable among the psychological factors; it is also the second most important variable in the model. Results show that punishment odds increase by 145.75% for every increase in hostility score. Age is the most critical variable out of the demographic factors, as the age increases by one year, the punishment odds increase by 16.84%.

Finally, all the clinical variables—SLE remission and glucocorticoids consumption—fall into the top five of the most relevant variables. For a lupus individual in remission, the odds of punishment rise by 12398.06%. Moreover, the consumption of each milligram of glucocorticoids increases the odds of punishment by 7.74%.

In the case of temporal decisions (Table 4), the most indispensable predicting variables came from psychological, clinical, decision-task, and social categories. In the psychological category, results show that anxiety, and paranoid ideation decrease the immediate reward odds by 60.16% and 67.2%, respectively, when they increase their score by one point. On the other hand, obsessive-compulsive, and depression increase the immediate reward odds by 97.08% and 50.8%, respectively, when they increase their score by one point. It is worth mentioning that anxiety and obsessive-compulsiveness are the two most influential variables in the whole model. Concerning clinical factors, both, remission and glucocorticoid consumption, fall into the most influential variables. On the one hand, Our approach was a within-subjects experimental design. So, once the individuals completed all the questionnaires, they participated in two standard behavioral-economic tasks. The first task was a third-party dictator game, focused on evaluating social punishment behavior in people with SLE in remission increase the immediate reward odds by 5148426.03%; on the other hand, those patients taking glucocorticoids increase their preference for immediate reward by 3136.55%. Regarding the decision task parameters, as the gaining rate increases by one unit, the odds for immediate reward decrease by 27.23%, as expected. Finally, among social variables, quality of life is the most relevant, as the quality of life score increases by one point, the odds for immediate rewards decreases by 27.23%.

## Discussion

In accordance with what was previously reported in the literature (see below), intrinsic task parameters, demographics, and psychological factors (excepting anxiety) behave as expected. To our knowledge, our work is the first one studying how lupus clinical factors affect social and temporal decisions.

Regarding intrinsic parameters from both tasks, i.e., personal cost in social decisions, and gaining rate in temporal decisions, both had the expected effect on participants’ responses. Our results are consistent with those reported in the literature on healthy populations: when personal cost increases, the odds of punishment decrease (Egas & Riedl, 2008; Ostrom et al., 1992); and when the daily gaining rate is higher, people behave patiently and prefer bigger delayed rewards (Amasino et al., 2019); (Berns et al., 2007). Referring to demographic factors, our results also behave as expected for healthy people. With age, people engage in more punitive behavior (Lee & Warneken, 2022);(Singh et al., 2014).

Concerning psychological factors, most of our results are consistent with previous results. Individuals with higher hostility scores are more likely to punish norm-violators (Raihani & Bell, 2019; Weng et al., 2015). Moreover, people with higher obsessive-compulsive scores, act impulsively and prefer sooner but smaller rewards (Ong et al., 2018). However, some studies have reported that higher anxiety scores are associated with a preference for immediate rewards; for instance, (Campbell & Egede, 2024) studied a diabetic population, and found that individuals who prefer immediate rewards had significantly higher symptoms of anxiety.We found the opposite; our results indicate that most anxious people act patiently, and prefer delayed but bigger rewards. These results are consistent with those reported by (Steinglass & Walsh, 2016), who studied populations with anorexia nervosa, obsessive-compulsive disorder, social anxiety disorder, and also a healthy population. They found that, across all samples, more anxious people acted patiently and showed greater preference for delayed but bigger rewards. So, additional research is needed to confirm what the most plausible effect of anxiety on decision-making is in people with SLE.

Finally, over all variables, remission in Our approach was a within-subjects experimental design. So, once the individuals completed all the questionnaires, they participated in two standard behavioral-economic tasks. The first task was a third-party dictator game, focused on evaluating social punishment behavior in people with SLE had the biggest effects on both, social and temporal decision-making. It is closely related to glucocorticoids consumption because glucocorticoids are the standard treatment to reach remission. In fact, people in remission and taking glucocorticoids showed greater odds of punishing unfair behavior, and also showed preference for immediate rewards.

It is possible that people in SLE remission had a history of chronic exposure to glucocorticoids, which could induce suboptimal decision-making, as demonstrated in mice by Cabeza et al., (2021).

However, we hypothesize that those collateral effects could also be observed in the general population, as reported by Putman et al., (2010), who administered 40 mg of glucocorticoids (cortisol) to healthy people, reporting a potential effect of glucocorticoids on the reward’s system.

We argue that, since many people around the world regularly take glucocorticoids to treat their seasonal allergic symptoms, it is necessary to conduct more investigation to separate the effects of chronic consumption from seasonal consumption.

Regarding limitations of our study, we didn’t control for how long our participants had taken glucocorticoids; as we only registered the current dosage in mg.

Despite our limitations, our study is a pioneer in investigating decision-making in people with SLE; it showed statistical power, and the consistency of psychological, demographic, social, and clinical variables, was validated with Mexican lupus datasets (see Ana Laura et al., 2023). Further studies must control the accumulated doses of glucocorticoids.

## Conclusions

This study was aimed at exploring how social, clinical, psychological, and demographic factors impact social and temporal decision-making in people with SLE. Our approach was a within-subjects experimental design. So, once the individuals completed all the questionnaires, they participated in two standard behavioral-economic tasks. The first task was a third-party dictator game, focused on evaluating social punishment behavior in people with SLE. For social decisions, we used the third-party dictator game, whereas, for temporal decisions, we used a delay-discounting task.

Concerning social decision, hostility, and age were essential predictors; for temporal decisions, anxiety and obsessive-compulsive figured as the most important. However, In both cases, clinical factors were critical decision-predictors. Even more influential than some intrinsic factors like punishment size or gaining rate. Firstly, when people with SLE are in remission, they tend to infringe higher punishment on those who violate the social norm, and they also tend to prefer immediate rewards instead of bigger but delayed rewards. Secondly, when people with SLE are taking glucocorticoids, they also prefer immediate rewards, and as the dosage of glucocorticoids intake increases, they tend to impose higher punishment on norm violators as well.

These results must be considered by clinicians, when they prescribe glucocorticoids to people with SLE, it could potentially affect decision-making. And by behavioral economics practitioners and researchers who should consider the effects of using synthetic corticosteroids in our current knowledge on decision-making.

## Acknowledgments

This work received support from Luis Aguilar, Alejandro León, and Jair García of the Laboratorio Nacional de Visualización Científica Avanzada. We also thank Carina Uribe Díaz, Christian Molina-Aguilar, and Alejandra Castillo Carbajal for their technical support. Authors would like to express their special acknowledgment to Fundación Proayuda Lupus Morelos A.C, Lupus MX, El despertar de la Mariposa, and Centro de Estudios Transdisciplinarios Athié-Calleja por los Derechos de las Personas con Lupus A. C., for their invaluable support.

## Data availability

Data is available on https://zenodo.org/records/10806272. All the analyses were developed and implemented in R version 4.3.3 (2024-02-29), and they are available on: https://github.com/NeuroGenomicsMX/Factors_affecting_decisions_in_SLE.

